# Cognition in younger women with premature ovarian insufficiency

**DOI:** 10.64898/2026.07.14.26358044

**Authors:** Laura F Naysmith, Laila Rida, Adam Hampshire

## Abstract

Premature ovarian insufficiency (POI) significantly impacts quality of life, yet the immediate cognitive landscape and lived experience of younger women remain under-researched. In 125 young women (aged 19-48; 66 with idiopathic POI, 59 age-matched controls), we examined self-reported cognitive distress and symptom burden within the POI cohort and compared objective global and domain-specific cognitive performance between groups. Objective accuracy scores were derived from six online tasks (‘Cognitron’) and combined into a robust global measure. Within the POI cohort, there were significant differences in symptom burden domains (χ²(3) = 61.90, *p*<0.001), with psychological and sexual symptoms reported at a significantly higher intensity than physical and vasomotor symptoms (all *p*<0.001). Furthermore, the standardised magnitude of perceived cognitive distress (56.20%) was significantly greater than that of overall symptom burden (42.00%, *p*<0.001). Case-control comparisons revealed no significant differences in global cognitive performance (*p*=0.615), yet the POI cohort performed significantly less accurate than controls in verbal analogical reasoning (–0.86 SD, 95% CI: –1.52, –0.20, p = 0.011). The findings highlight an urgent need for comprehensive emotional and psychosexual support in POI care. Additionally, the presence of high cognitive distress alongside localised objective deficits demonstrates that cognitive health monitoring must be proactive in early adulthood, especially given their established long-term risks for later-life cognitive decline and dementia.

## Introduction

Premature Ovarian Insufficiency (POI) is more than a reproductive disorder. Women with POI are not only at increased risk of cardiovascular disease^1–3^ and osteoporosis^4–7^, but also of depression^8^, anxiety^9^, poor life quality^10^, negative psychosocial impact^11^, and suicidality^12^. Moreover, there is increasing evidence of both cognitive distress^12,13^ and neurological risk, such as dementia^14,15^.

POI is the loss of ovarian activity at, or under, age 40 resulting in a chronic hypoestrogenic state. Oestrogen is typically regulated centrally via the hypothalamic-pituitary-ovary (HPO) axis and act widely within the brain to support neuroprotective, neurotrophic, metabolic, and neurotransmitter-modulating processes. Therefore, hypoestrogenism may be associated with changes in neural structure and function^16^. Evidence has primarily come from the menopausal transition showing age-independent changes in neural structure and metabolism^17–19^, in addition to connectivity^20,21^. Although sex steroid hormone changes may be key to understanding brain health in menopause, there are other factors that are also emerging, such as vasomotor symptoms^22,23^, type of menopause^24,25^, hormone treatment^26^, and, associated with POI, the age of menopause^27,28^.

Emerging evidence has shown early structural alterations in the brain in women with idiopathic POI^15^, in addition to associations to increased risk of dementia^14^. Moreover, younger age at menopause has been associated with cognitive impairment^29,30^. Objectively measured cognition provides insight into brain function and utilising these task measures of accuracy or reaction time on various cognitive domains can identify subtle differences in brain function. Investigating the influence of POI and earlier age at menopause on later life cognitive function has previously been examined, with research illustrating greater risk for cognitive deficits at later life^30–32^. Yet, whether we see subtle changes in cognitive deficits at a younger age in women diagnosed with POI is unclear, indeed how symptoms interplay with cognitive function may be important and provide essential evidence of whether cognition, as a proxy of brain function, may show deficits which may be associated with POI. This approach will provide more understand of POI and neurological health without the confounding effects of age-related comorbidities.

With a validated online platform for cognitive testing (“Cognitron”)^33^, younger women with POI completed six cognitive tasks, in addition to the Greene Climacteric Scale^34^ to measure symptom burden and MENO-COG^35^ to measure cognitive distress. Firstly, given the profound psychosocial impact associated with POI^11^, it is hypothesised that the psychological domain of the Greene Climacteric Scale will exhibit significantly higher typical severity scores (item-averaged means) compared to all other domains within the POI cohort. Furthermore, the POI cohort will demonstrate greater standardised cognitive distress compared to total general symptom burden due to the high prevalence of these symptoms and associated neurological vulnerability associated with changes to HPO axis.^36^ Comparing the two cohorts, the POI cohort are expected to perform less accurately both globally and task-specifically, than age-matched controls. They were also expected to show more intra-individual cognitive variability to age-matched controls, which may be associated with hypoestrogenic symptoms. Menopause research suggests that cognition becomes more variable due to the interactions between vasomotor symptoms, sleep disruption, mood, lifestyle factors which may ameliorate cognitive variability, thus intra-individual cognitive variability may provide meaningful insight into brain function.^37^ Subjective measures of symptom burden and cognitive distress were expected to correlate negatively with both global and task accuracy scores, suggesting greater cognitive deficits with more symptom distress.

## Methods

This study received ethical approval from the King’s College London Research Ethics Committee (HR-24/25-51594).

### Participants

Women with POI were recruited through online posters disseminated via open-access social media platforms, the Daisy Network charity, and King’s College London recruitment circulars between October 2025 and January 2026. Participants were eligible if they self-reported a diagnosis of POI from a medical professional at or before age 40, were aged 18 years or older, and did not have a current neurological or psychiatric diagnosis. Information on diagnostic investigations and aetiology was collected for descriptive purposes only.

Normative data from age-matched controls came from the Great British Intelligence Test (GBIT), a nationwide study conducted in the United Kingdom (https://gbit.cognitron.co.uk). GBIT received ethical approval from the Imperial College Ethics Committee (REC Ref ICREC/SETREC 17IC4009). Recruitment procedures and cohort characteristics are comprehensively described by Balaet et al.^38^ The initial GBIT cohort included 346,780 adults aged over 18 who completed the baseline assessment in 2020. Between August and September 2025, 246,569 participants were invited to complete a follow-up survey and cognitive assessments. This analysis included a subset of individuals who completed the same six cognitive tasks as the POI cohort and provided complete survey data on menopausal status and HT use. To create an age-matched control group, a subset of GBIT participants was selected using a propensity score matching approach (see Sample Characteristics).

### Survey

Data from the POI cohort were collected with a self-administered online survey using the King’s College London Qualtrics platform. All participants who provided informed consented were presented with the survey questions. POI diagnosis was self-reported as either; i) unknown (idiopathic), ii) related to medical treatment or surgery, iii) confirmed diagnosis of an autoimmune condition, or iv) confirmed diagnosis of a known genetic cause. Alternatively, participants could report “unsure”. No independent verification of autoimmune or genetic testing was performed.

Current HT use was assessed via self-report. Participants who responded “Yes” to current use were classified as HT users, while those responding “No” were classified as non-users. HT users provided length of current use which ranged from “Less than 6 months” to “2+ years” and participants could select all HT types they currently used (Figure 1).

**Figure 1.**
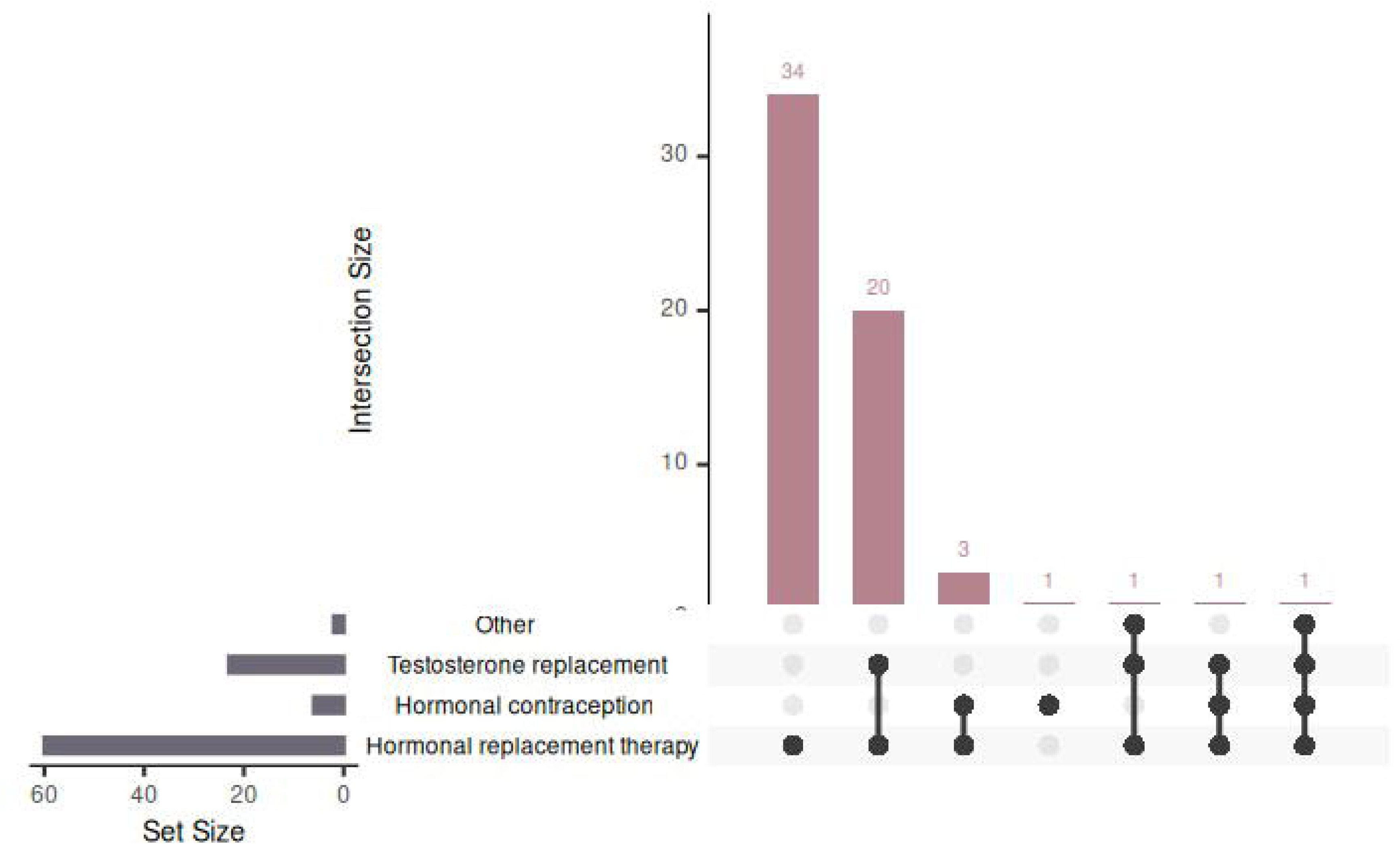
UpSet plot visualising current hormone treatment (HT) use in 61 POI cohort.

The Greene Climacteric Questionnaire^34^ was used to assess current symptom burden. Participants were asked to indicate “the extent to which you are bothered at the moment by any of these symptoms” rating from 1=not at all, 2= a little, 3= quite a bit, 4=extremely. To facilitate comparisons across domains, composite subscale scores were created and then standardised using z-scores, calculated by subtracting the sample mean and dividing by the standard deviation for each composite score. Based on the Greene Climacteric Scale^34^, symptoms were combined to form composite scores representing physical, psychological, vasomotor, and sexual domains, in addition to a total symptom burden score. These composite scores reflect current symptom burden, with higher scores indicating greater severity.

The MENO-COG Scale^35^ was used to assess perceived cognitive distress. From the past two weeks, participants rated their experience as 1=never, 2=rarely, 3=sometimes, 4=often, or 5=all the time. A total summary score was created and standardised using z-scores, calculated by subtracting the sample mean and dividing by the standard deviation for each score. A higher composite score reflects greater perceived cognitive difficulty over the past two weeks.

### Assessment of objective cognitive performance

For the POI cohort, participants completed six computerised tasks on the Cognitron platform within one hour of completing the survey. The six tasks were as per Hampshire et al.^33^ (see Supplementary Materials). Tasks were presented to participants in a fixed order and were accessed via personal devices, such as laptop computer or smartphone. The tasks were selected from the broader Cognitron library to be decorrelated, thus, each distinct task captures a cognitive domain, while still providing robust global cognitive composite scores^39–41^. They included measures of immediate and delayed memory recall, 2D manipulation, block designs, verbal analogical reasoning, and switching Stroop interference. Each task outputs a primary accuracy-based score and secondary reaction times and error types. This manuscript primarily focuses on accuracy based composite scores. The raw, unadjusted data were processed by applying a rank-based transformation to normality, outputting z-scores for each participant on each task metric. Global cognitive accuracy scores were calculated as the standardised (z-score) mean of the standardised accuracy.

Intra-individual cognitive variability, a within-person cognitive variability score was calculated for each participant by computing the standard deviation across their z-scored cognitive task scores to yield a single metric. To facilitate comparison and interpretation in regression analyses, the variability index was standardised.

### Statistical analysis

POI and GBIT data were processed and analyzed in Jamovi version 2.7.7 and R Studio 4.5. In the POI cohort, all survey responses and cognitive task responses were reviewed for completeness. For the survey, no outliers were excluded, as extreme responses reflected clinically meaningful symptom experiences. Missing data were handled listwise, with participants excluded only from analyses where relevant data were missing. For the online cognitive battery, data cleaning and quality control focused primarily on response time distributions to ensure the validity of task scores. No participants were excluded based on accuracy, as task accuracy summary scores remained within expected ranges; however, outliers were retained to allow for task-specific sensitivity analyses. To ensure scores reflected realistic cognitive engagement, performance data were converted to missing values if a participant’s median reaction time for each task met predefined exclusion criteria. This included a cutoff of ≤ 500ms on the 2D manipulations, verbal analogies, switching Stroop interference, and Block design tasks. The number of times a participant left their browser window while completing a task was set to exclude anyone who did this more than once.

### Sample characteristics

Demographic characteristics of the POI and age-matched cohorts are included in Table 1. HT use in POI is visualised in Figure 1. Differences in age and HT use were compared across POI and age-matched control groups, using independent *t*-tests and a chi squares test of independence, respectively.

**Table 1.** Demographics of cohort.

| Group, n | POI, 66 | Age-matched, 59 |
| --- | --- | --- |
| Mean age (SD; age range) | 35.60 (7.42; 19.00-48.00) | 35.70 (7.14; 19.00-46.00) |
| Mean age at diagnosis (SD; age range) | 29.0 (7.93; 15.00-39.00) | - |
| Etiology of POI, % cohort |  |  |
| Idiopathic | 91.40 | - |
| Autoimmune disease | 2.90 | - |
| Genetic condition | 5.70 | - |
| Ethnicity, % cohort |  |  |
| Asian | 2.90 | 9.20 |
| Black | 1.40 | 1.70 |
| White | 90.00 | 62.70 |
| Mixed | 2.90 | 3.40 |
| Not Stated | 2.80 | 23.00 |
| Current educational attainment, % cohort |  |  |
| Secondary school (GCSE, A Level) | 25.70 | 22.00 |
| Undergraduate degree | 31.40 | 50.90 |
| Postgraduate degree | 42.90 | 5.10 |
| Not Stated | 0.00 | 22.00 |
| Lifestyle factors, % cohort |  |  |
| Current smoker | 15.70 | - |
| Excessive alcohol consumption | 5.70 | 15.30 |
| Current hormonal treatment (HT) use, % cohort<sup>1</sup> | 94.20 | 6.80 |
| Mean age for HT initiation<br>(SD; age range) in current HT<br>users | 30.70 (7.89; 15.00-44.00) | - |
| Duration of current HT use, %<br>of current HT users |  |  |
| Less than 6 months | 18.20 | 1.70 |
| 6-12 months | 15.10 | 0.00 |
| 1-2 years | 25.80 | 1.70 |
| 2+ years | 40.90 | 3.40 |
| Not Stated | 0.00 | 93.20 |

<sup>1</sup> See Figure 1 for UpSet plot of distribution of current HT. Excessive alcohol consumption classified as more than 14 units per week.

To ensure comparability between the cohorts, a propensity score matching procedure was implemented. From the whole GBIT sample, a normative pool was first filtered to include only female participants with complete data for the primary variables of interest: age and six task accuracy scores. Propensity scores, representing the probability of a participant belonging to the experimental group, were estimated using a logistic regression model with age as the primary predictor. Using these scores, participants were matched via a 1:1 nearest-neighbour algorithm without replacement, where each experimental participant was paired with the control from the normative pool who possessed the closest estimated propensity.

### POI symptoms

For both the Greene Climacteric Scale and MENO-COG scale, proportional means and item-averaged medians were derived by dividing the raw summary scores by the number of items within each respective subscale (Table 2). Thus, rescaling the symptom scales back to the original Likert units (e.g., 0–3), representing the “typical” severity reported per domain. Shapiro-Wilk and skewness are reported to assess how symptoms are experienced across the cohort. For the Greene Climacteric Scale domains, a Friedman Test was conducted using the item-averaged (proportional) mean scores. Significant main effects were further investigated using Durbin-Conover pairwise comparisons to identify specific differences in symptom severity between domains.

**Table 2.** Summary statistics for symptom burden and perceived cognitive distress.

| Summary score | Raw Median [IQR] | Proportional Mean | Item-Averaged Median | POMP, % (SD) | Shapiro-Wilk ($p$-value) | Skewness (SE) |
| --- | --- | --- | --- | --- | --- | --- |
| Current symptom burden from 4-point Likert scale [0-3] |  |  |  |  |  |  |
| Total | 26.00[12.80-21.00] | 1.26 | 1.24 | 42.00 (16.20) | 0.98 (0.498) | 0.34 (0.287) |
| Psychological burden | 17.00[8.75-11.00] | 1.54 | 1.55 | - | 0.99 (0.881) | -0.01 (0.287) |
| Physical burden | 5.00 [5.00-7.00] | 0.87 | 0.71 | - | 0.94<br>(0.001) | 0.86<br>(0.287) |
| Vasomotor burden | 1.00 [2.75-2.00] | 0.78 | 0.50 | - | 0.82<br>($<0.001$) | 1.04<br>(0.287) |
| Sexual burden | 2.00 [2.00-1.00] | 1.79 | 2.00 | - | 0.85<br>($<0.001$) | -0.28<br>(0.287) |
| Cognitive distress from 5-point Likert scale [0-4] |  |  |  |  |  |  |
| Total | 65.00 [53.30-84.80] | 2.25 | 2.46 | 56.20<br>(19.90) | 0.977<br>(0.224) | -0.318<br>(0.287) |

IQR= Interquartile range; POMP= Percentage of Maximum Score Possible; SD= Standard deviation; SE= Standard error

A non-parametric Wilcoxon Rank test was used to compare the percentage of maximum possible (POMP) due to non-normal data distribution. The POMP provides a standardised comparison of the two scales with different numbers of items and Likert scale to assess differences in symptom experience. A Pearson correlation was conducted to assess the relationship of symptoms.

### Objective cognitive performance

A linear regression model was conducted to measure the relationship between global cognitive accuracy and separately for cognitive intra-individual variability, using the same predictor variables. For both models, standardised coefficients were calculated indicating the difference in global cognitive accuracy/cognitive intra-individual variability for a one standard deviation change based on group. Details of the model outputs, including estimates and significance, are provided in the Results section for group and HT main effects, education, and ethnicity are reported.

Summary scores for each of the six cognitive tasks were analysed with linear regression models. Details of the model outputs, including estimates and significance, are provided in the Results for group and HT main effects, education, and ethnicity are reported.

### Associations between POI symptoms and objective cognitive performance

For the POI cohort, Pearson’s correlation was used to determine the relationship between total symptom burden, total perceived cognitive difficulty, global cognitive accuracy, cognitive variability, and summary scores for each task. Correlation matrices are presented in Table 3.

**Table 3.** Spearman’s rank (r) correlation matrix between total symptom burden, cognitive distress and global cognitive accuracy, cognitive variability, and cognitive domain-specific task accuracy. *p<0.05.

| Item | Global<br>Cognitive<br>accuracy | Cognitive<br>Variability | Block<br>Design | 2D manipulation | Delayed<br>memory<br>recognition | Immediate<br>memory<br>recognition | Stroop<br>interference | Verbal<br>analogical<br>reasoning |
| --- | --- | --- | --- | --- | --- | --- | --- | --- |
| Total<br>symptom<br>burden | -0.21 | 0.03 | -0.14 | -0.15 | -0.03 | -0.16 | -0.10 | -0.24 |
| Cognitive<br>distress | -0.15 | 0.19 | -0.16 | -0.01 | -0.11 | -0.17 | 0.88 | -0.25* |

## Results

### Sample characteristics

In the POI cohort, 76 participants consented to the study, all meeting the study inclusion criteria. Of the 76 participants, 75 completed the survey and one was removed as an incomplete duplicate event. Two participants were removed for indicating “Not sure” regarding their POI diagnosis. Due to known influence of surgical menopause on cognition and symptom presentation, we excluded three individuals from the analysis who reported surgical POI. Three participants were removed after cleaning their cognitive data, leaving a final analytic sample of 66.

For the age-matched controls, using the 1:1 nearest neighbor algorithm without replacement, we identified 66 women from the GBIT cohort with full cognitive data to match the final POI cohort. We excluded 1 woman who was postmenopausal and two who had hysterectomies. Four women had missing HT data. The final analytic sample was 59.

The mean age of the POI cohort was 35.60 years (SD=7.42) and in the age-matched controls was 35.70 years (SD=7.14). Age did not differ significantly (Welch’s *t*(123)=0.003, *p=*0·976). HT use varied by cohort (χ²(1)= 91.50, *p<*0·001) with standardised residuals reported in full in the Supplementary Materials. All demographic information is reported in Table 1 and Figure 1 visualizes current hormonal treatment (HT) within the POI cohort.

### POI symptoms

Table 2 shows summary statistics for symptom burden and perceived cognitive distress. While psychological and total burden followed a normal distribution, the physical, vasomotor, and sexual subscales were significantly non-normal (all *p*<0.001). Specifically, vasomotor symptoms exhibited a high positive skew, indicating a clustering of low-severity reports.

A Friedman test revealed a significant effect of symptom domain on reported symptom burden, (χ² (3)=61.90, *p*<0.001). Post-hoc Durbin-Conover pairwise comparisons indicated that psychological and sexual symptoms were reported at a significantly higher intensity than physical and vasomotor symptoms (all *p*<0.001). Notably, no significant difference was found between psychological and sexual symptom intensity (*p*=0.783), nor between physical and vasomotor intensity (*p*=0.610).

A Wilcoxon Rank showed significant differences between total current symptom burden and total perceived cognitive distress (*p*<0.001). On a standardised 0–100 scale, the magnitude of perceived cognitive distress (56.20%) was significantly greater than the magnitude of overall symptom burden (42.00%) in POI. There was a significant positive association between symptom scales, with women experiencing greater perceived cognitive difficulties over the past two weeks also showing higher current symptom burden (r=0.480, *p*<0.001).

### Objective cognitive performance

No significant differences were observed between the POI and age-matched controls for global cognitive performance (*p=*0.615) or intra-individual cognitive variability (*p=*0.198) (Figure 2). However, we observed a main effect of HT on global cognitive performance, with women on HT performing worse than women not on HT (–0.79 SD, CI 95%: –1.54, –0.04, *p=*0.038). There was no main effect of HT on intra-individual cognitive variability (*p=*0.556). Moreover, in both models there was no main effect of education or ethnicity (*p*>0.050).

**Figure 2.**
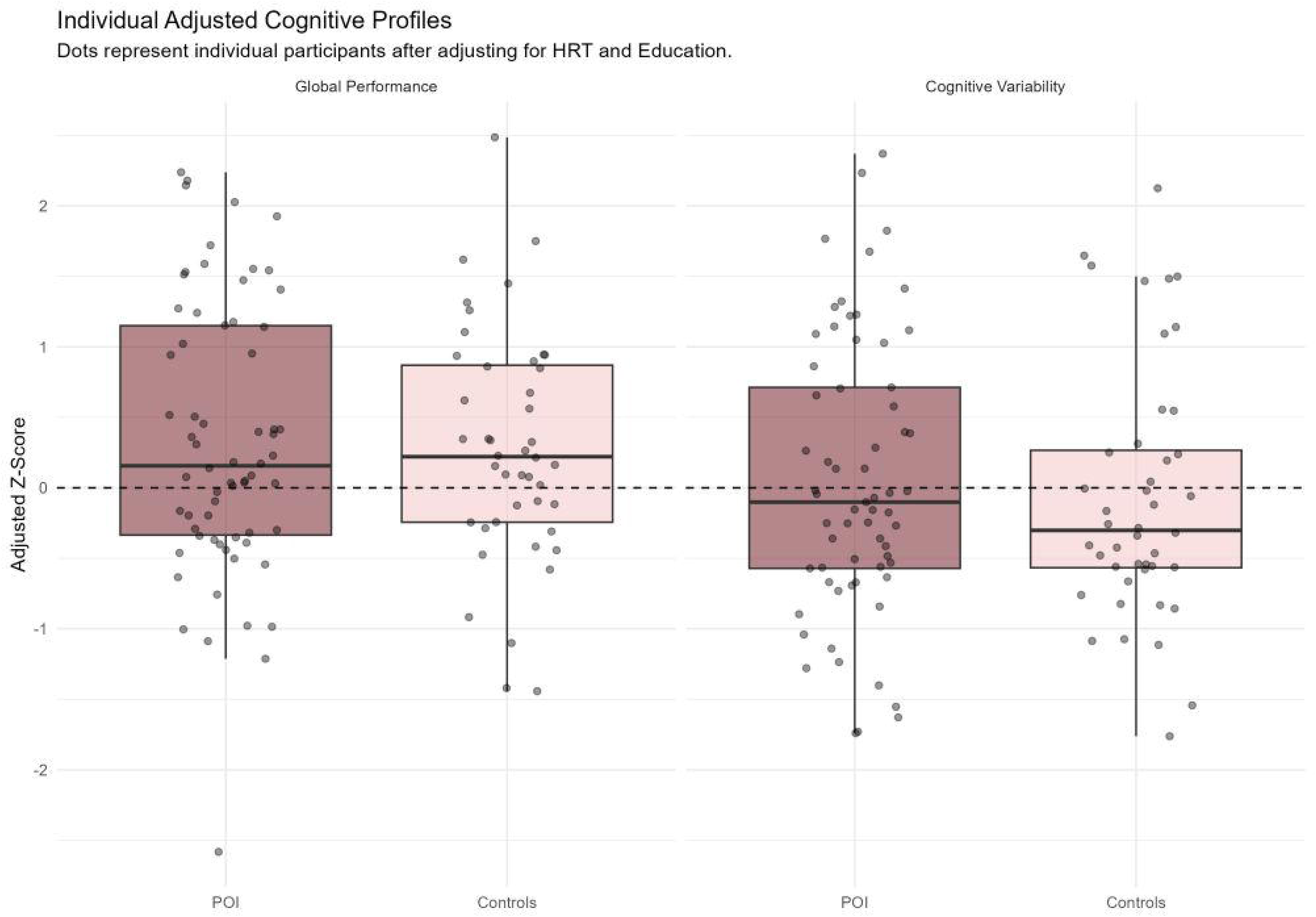
Global cognitive performance and cognitive variability in POI and age-matched controls. Boxplots represent the distribution of adjusted z-scores. Adjusted z-scores are standardised residuals derived from linear regression models. The horiontal line represents the sample mean.

Figure 3 and Figure 4 visualises performance across tasks and groups. There was a significant main effect of group on the verbal analogical reasoning task (–0.86 SD, CI 95%: –1.52, –0·20, *p=*0.011), with the POI cohort performing less accurately than the age-matched controls. However, there were no significant differences in performance between groups on the 2D manipulation (*p=*0.113), block design (*p=*0.452), immediate memory recognition (*p=*0.808), delayed memory recognition (*p=*0.473), or Stroop interference (*p=*0.399) tasks.

**Figure 3.**
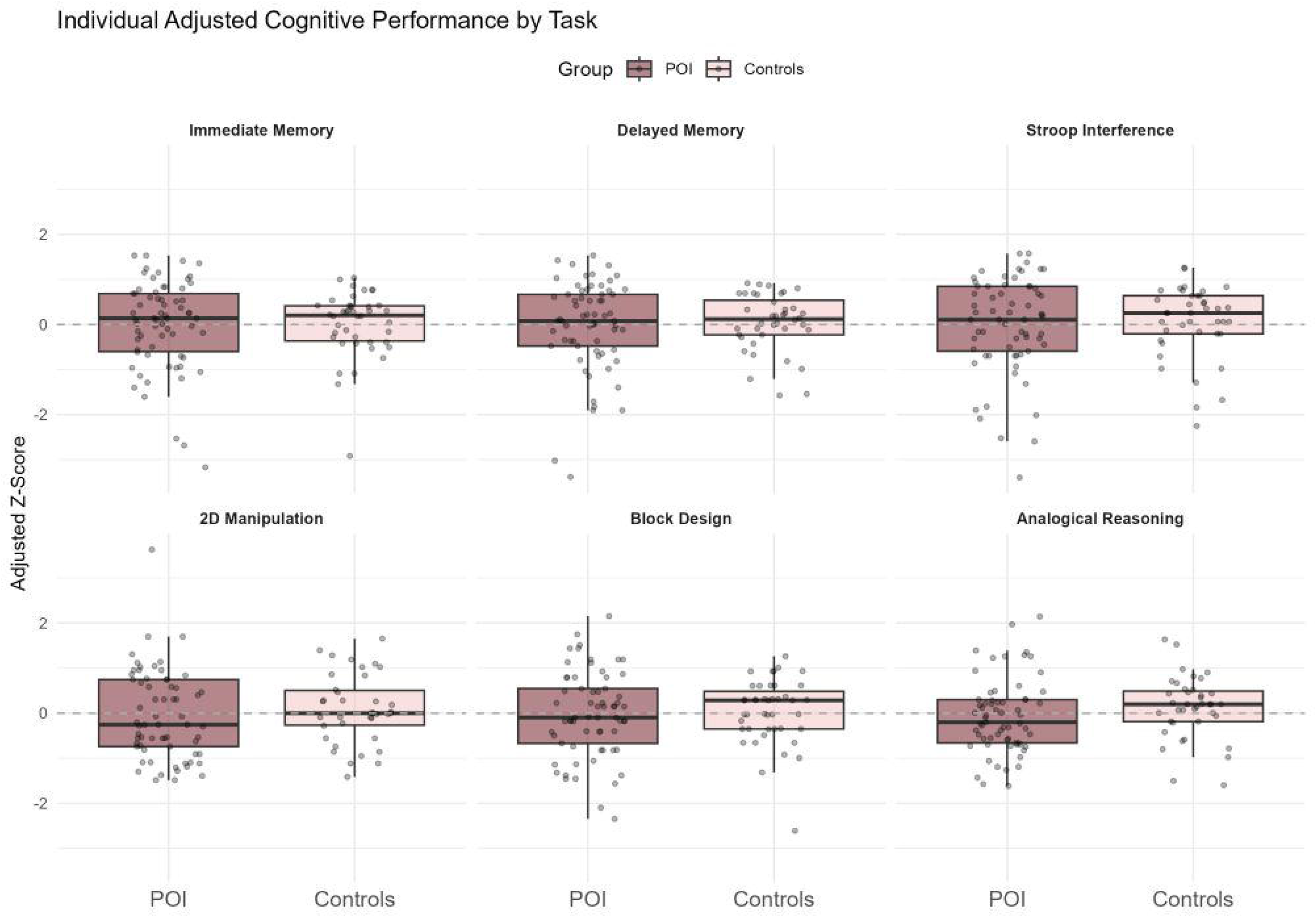
Task performance on six cognitive tasks in POI and age-matched controls using boxplots. This represents the distribution of adjusted z-scores. Adjusted z-scores are standardised residuals derived from linear regression models. The horizontal line at represents the sample mean.

**Figure 4.**
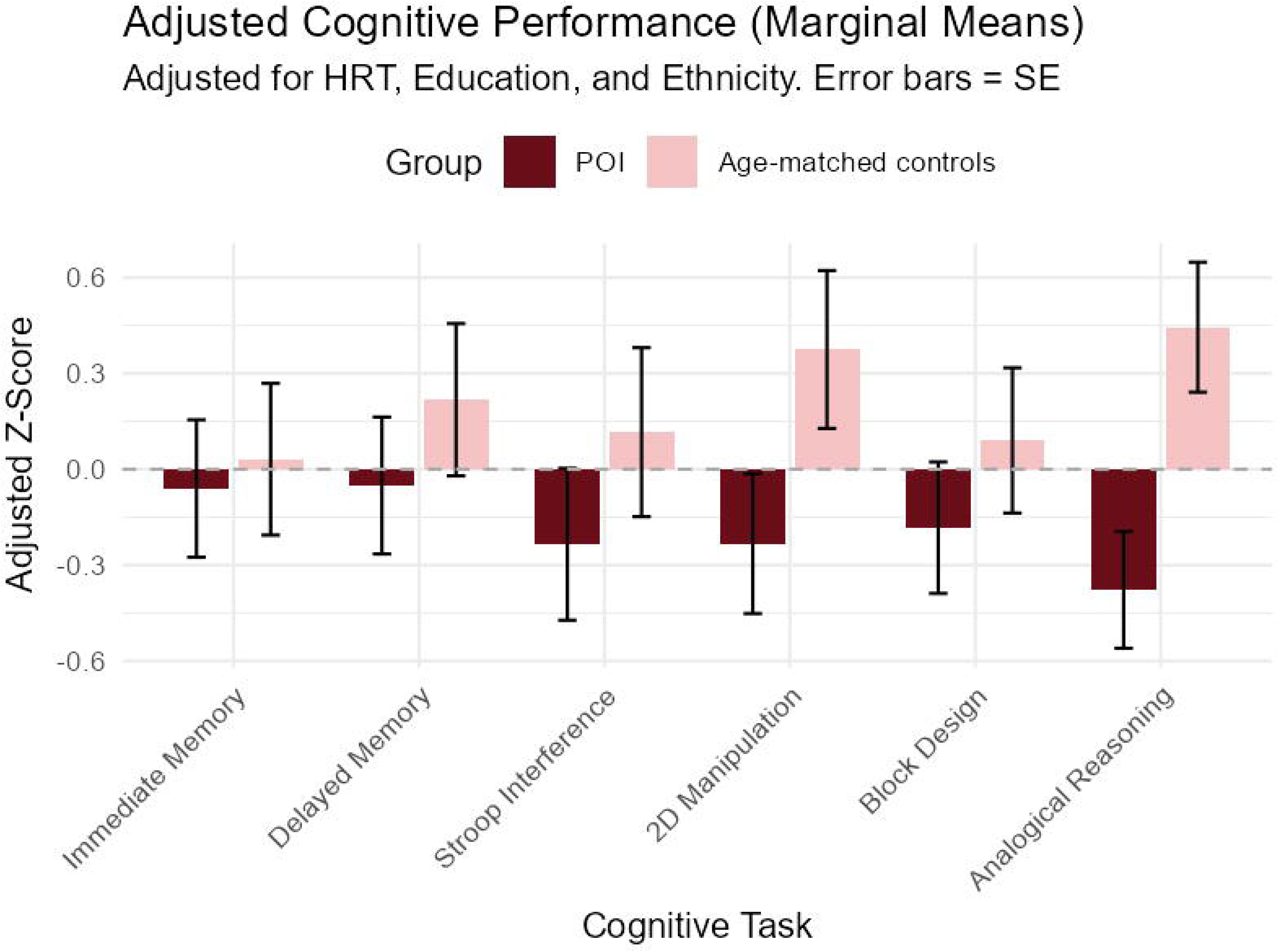
Task performance on six cognitive tasks in POI and age-matched controls using bar charts. This represents the group mean adjusted z-scores across six cognitive domains. Error bars represent the standard error of the mean. Adjusted z-scores are standardised residuals derived from linear regression models.

There was a main effect of HT on delayed memory recognition (–0.75 SD, CI 95%: –1.45, – 0·06, *p=*0.033) and immediate memory recognition (–0.88 SD, CI 95%: –1.57, –0·17, *p=*0.015) tasks. HT users performed less accurately than non-users, however, there were no significant differences in performance between groups on the Stroop interference (*p=*0.705), block design (*p=*0.084), verbal analogical reasoning (*p=*0.386), 2D manipulation (*p=*0.790) tasks.

On the verbal analogical reasoning task, there was a main effect of education (0.55 SD, CI 95%: 0.16, 0.94, *p=*0.006) with more accurate performance in those educated with a university degree, compared to secondary school education. No other main effects of education were observed on the remaining tasks (all *p*>0.050). No main effect of ethnicity was observed on any task (all *p*>0.050).

### Associations between symptoms and objective global cognitive performance

There was no association between global cognitive performance with total symptom burden (r=-0.205, *p*=0.098) or perceived cognitive distress (r=-0.151, *p*=0.225), nor was there an association between global intra-individual cognitive variability with total symptom burden (r=0.033, *p*=0.794) or perceived cognitive distress (r=0.188, *p*=0.133).

A weak negative correlation was observed between perceived cognitive distress and the verbal analogical reasoning task (r=-0.205, *p*=0.044), but no other significant correlations were observed (all *p*>0.050). The full correlation matrix can be found in Table 3.

## Discussion

In a cohort of younger women with idiopathic POI, we observed a distinct symptom profile whereby psychological and sexual symptom burden were significantly greater than vasomotor and physical symptom burden. Moreover, the POI cohort reported a level of perceived cognitive distress that significantly exceeded their total standard symptom burden. When examining objectively measured cognition against age-matched controls, we observed no significant differences in global cognitive accuracy or intra-individual cognitive variability. However, a significant, domain-specific deficit with a large effect size emerged in verbal analogical reasoning. Interestingly, perceived cognitive distress was only significantly associated with this verbal analogical reasoning task, showing no relationship with other objective cognitive outcomes. This clear divergence underscores that standard global cognitive screens may fail to capture the distressing everyday cognitive burden reported by younger women with POI.

Whilst the clinical presentation of POI symptoms is heterogenous,^42^ our findings offer clear evidence into the lived experience of younger women, demonstrating that psychological and sexual distress heavily dominate their symptom profile. POI is associated with a three– and four-fold increased risk of depression and anxiety^9^ and poorer quality of life.^43^ Sexual dysfunction in POI has been identified as multi-dimensional, with the combination of early genitourinary symptoms and lower Female Sexual Function Index.^44^ Taken together, this distinct clinical profile underscores an urgent need for POI care to integrate targeted emotional support and counselling. The elevated risk for long-term mental illness and diminished quality of life highlights a critical care gap. Managing the complex and heavy burden of these psychosocial and psychosexual symptoms requires a holistic approach combining both hormonal and non-hormonal strategies^42^.

Next, our findings further illustrate that perceived cognitive distress is a major, yet frequently overlooked, component of the lived experience of younger women with POI. Driven by changes in the HPO axis, cognitive symptoms are common across various reproductive milestones and ovarian disorders, yet they remain severely under-researched in the context of POI.^36^ Moreover, identifying this high cognitive burden at a younger age is crucial as it offers a window for proactive, preventative support rather than reactive care. For example, we know that POI is associated with increased risk for dementia^14^ and early evidence for neural alterations associated with idiopathic POI who are HT naieve.^15^ In addition, our findings also raises questions about bout how symptoms are clinically measured in this population; widely used tools like the POIQOLS^45^ or the Greene Climacteric Scale^34^ omit perceived cognitive distress. Thus, future research should develop clinically valid measures which capture lived experience of symptoms in POI, including cognitive distress.

Interestingly, our evidence provides novel insight into how objectively measured cognition in POI presents in early adulthood, with domain-specific cognitive deficits rather than broad global cognitive deficits. While previous research has predominantly looked at POI through a critical lens of later-life dementia risk, our study captures the immediate cognitive landscape during the lived experience of the condition. Existing mid-to-late-life literature consistently reports more widespread deficits. Ryan et al.^30^ previously reported a two-fold increase in visual memory and verbal fluency impairments in an older cohort with POI (age M=74.10). In addition, there was 35% increased risk for global cognitive decline in women with POI over seven years. Nakanishi et al.^32^ observed persistent deficits in orientation, memory recall, and verbal fluency in women who underwent menopause before age 40 compared to those over 50, and Mensegere et al^31^ reported poorer global and task-specific cognition in an older cohort (age M=61.00) who experienced menopause before age 45. Our findings might suggest subtle and localised differences in cognition between younger women with POI and age-matched controls. This subtle shift in cognition may become more observable for the long-term effects of aging. As POI is associated with a 35% increased risk of global cognitive decline in later life, further research is warranted to better understand the cognitive trajectories in women at a younger age.

Moreover, cognitive distress was only significantly associated with verbal analogical reasoning accuracy, whereby women with greater perceived cognitive distress performed less accurately on this task. Cognitive distress did not correlate with any other objective cognitive measures, nor did symptom burden. Previously, in menopausal women, we observed that subjective cognitive symptoms like reporting brain fog and poorer memory were not strongly associated with objective cognitive performance.^46^ In the current study, this persistent disconnect may illustrate that standard objective measures fail to capture cognitive distress. However, verbal analogical reasoning, which evaluates fluid intelligence and complex relational thinking, may be uniquely sensitive to the subtle cognitive strain reported by younger women with POI, rendering it more susceptible to performance dips under conditions of high perceived distress.

Finally, it is important to note the HT finding observed in the current study, whereby on both memory recognition tasks, HT users performing significantly less accurate than HT non-users. Because our data are cross-sectional, this relationship must be interpreted with caution. Without detailed metrics on HT formulation, dosage, or long-term adherence, we cannot infer a causal link. Instead, this finding likely reflects a phenomenon of confounding by indication; younger women experiencing the most severe, debilitating POI symptoms or baseline cognitive changes may be the most likely to seek out and remain on HT. With limited randomised controlled trials (RCT) for hormone treatment on cognition, in both menopause and POI, the data are inconclusive about how HT may affect cognitive performance.^26^ Further research is required to address this in POI, and the benefits of HT use on neurological health is a critical gap in POI care support.

The current study has limitations. First, our cohort size is relatively small, meaning a larger sample would be required to ensure broader generalizability of the findings. However, it is notable that despite this sample size, a large effect size was still observed for the domain-specific deficit in verbal analogical reasoning. Second, while evaluating cognition in this younger cohort is a necessary first step toward understanding POI, longitudinal data are urgently needed to assess whether long-term cognitive trajectories differ between groups over time. Prospective tracking would also clarify whether fluctuations in the lived experience of symptoms are associated with these cognitive trajectories. Finally, clinical metrics included in this study (Greene Climacteric Scale; MENO-COG scale), were originally developed to assess menopausal symptoms in midlife. Our findings emphasise the differences in psychosocial impact of POI, such as infertility at a younger age and aging ovaries in a younger body^47^. Consequently, there is a pressing need to develop and validate specific tools designed to capture the unique lived experience of younger women with POI.

## Data Availability

All data produced in the present study are available upon reasonable request to the authors.

