## Supplementary Materials for "Cognition in younger women with premature ovarian insufficiency"

*Objective cognitive assessments*

The Cognitron Battery of tasks are detailed as per Hampshire et al., 2024.

***Immediate Memory*** - 20 images are displayed with an encoding time of 2000ms and inter stimulus interval of 500ms. After the full sequence is finished, each stimulus is then presented in turn within a grid of 7 distractors. The distractors are categorised by how similar they are to the stimulus along 3 dimensions. The dimensions are:

- Item - Whether the image is of the same item as the target.
- Precision - Whether the image is of the same pose or artistic style as the target. - Mis-binding - Whether the image is flipped left or right.

The main outcome measure is the number of points accrued, where points are awarded for each trial as incorrect item = 0, correct item = 1, correct item + precision = 2, correct item, precision and binding = 3, summed across trials. Secondary scores include error types and median response times.

***Delayed Memory*** - The volunteer is presented with the same task as the Object Memory (Immediate Memory) task described above, however the encoding period is removed. This means the volunteer must remember the stimuli from the beginning of the battery. The main outcome measure is the total number of points accrued as described in the Object Memory (Immediate Memory). Secondary scores include error types and median response times.

***2D Mental Manipulation*** - A target grid of coloured blocks is displayed above a set of 4 similar grids of colored blocks. 3 of the grids have been subtly altered from the target and the 4th is identical to the target. The grids have all been randomly oriented at 90, 180 or 270 degrees from the target grid. The volunteer must click the grid that is identical to the target as quickly and accurately as possible. They have 3 minutes to complete as many trials as they can. The main outcome measure is the total number of correctly identified matches divided by the total number of trials completed. Secondary score is median response time.

***Analogical Reasoning*** - The volunteer is presented with a set of comparative analogies, for example "light is to heavy as fast is to slow". The volunteer must work out if the comparison is correct. In the given example, this comparison is correct as they are both opposites of each other. An example of an incorrect analogical comparison would be "human is to ape as fast is to slow". The volunteer must complete as many comparisons as possible in 3 minutes as accurately as possible. The main outcome measure is the total number of correct trials divided by the total number of completed trials. Secondary score is median response time.

**Block Design** - Participants are presented with two 7x7 grids of squares displayed side-by-side. The left grid contains a configuration of coloured contiguous blocks, while the right grid displays a target shape made of a single black block. Participants must select the coloured blocks in the left panel to remove them so that the remaining structure perfectly matches the target shape on the right. A key rule is that blocks will fall under gravity if they are left unsupported from below. If a participant removes an incorrect block, the trial immediately ends. The assessment typically involves 12 to 15 trials, with difficulty progressively scaling based on the number of blocks that must be removed and how many blocks need to fall into an empty space to achieve the final target shape.

**Stroop Interference** - Participants are presented with the words "blue" or "red" printed in either blue or red ink, creating situations where the text and colour are either congruent or incongruent. A prompt box on the screen indicates the specific rule for that trial: participants must identify either the colour of the ink or read the text of the word. The relevant rule (colour vs. text) periodically switches throughout the assessment. Participants must tap the correct answer based on the current rule as quickly and accurately as possible, typically across 60 trials.

**Table S1.** Standardised residuals from chi square independence test on HT use based on group. * $p<0.05$

| | **Group** | |
|---|---|---|
| **HT use** | **POI** | **Age-matched controls** |
| **No** | **-9.57*** | **9.57*** |
| **Yes** | **9.57*** | **-9.57*** |
